# Melanonychia and use of nail biopsy to screen for subungual melanoma in a large ethnically diverse pediatric cohort: A cross-sectional study

**DOI:** 10.1101/2022.08.11.22278683

**Authors:** Helen H. Park, Alexander Lieu, Rosalynn R.Z. Conic, Lauren Metterle, Sijia Zhang, Isabella Toledo, George K. Hightower

## Abstract

**Background:** There are no widely accepted pediatric guidelines on who should undergo biopsy of the nail apparatus to screen for subungual melanoma (SUM). Reported cases of pre-adolescent children diagnosed with SUM presenting as melanonychia remain controversial. Further, observed age differences in the incidence of acral lentiginous melanoma are complicated by reported ethnic differences in adult prevalence and survival rates.

**Methods:** We conducted a retrospective chart review of Rady Children’s Hospital San Diego, a tertiary hospital system, with over 2 million patients in its electronic health records to identify patients diagnosed with melanonychia younger than 12 years and biopsies in these children, between January 1, 2010, and July 31, 2021.

**Results:** There were 623,805 patients younger than 12 years of age seen in the outpatient setting. Average age was 7.2 years for melanonychia diagnosis, and 5.1 years for those biopsied. Nail apparatus biopsies were performed in 22 different individuals and involved the hand 17/22 (77%) and the foot 5/22 (23%). The presence of Hutchinson’s sign was documented in 9/22 (41%) patients. Males were diagnosed with melanonychia and underwent biopsy more often than females. Many of the biopsies were performed in Hispanic/Latino 11/22 (50%) and Asian/Pacific Islander 3/22 (14%) patients.

**Discussion:** Regardless of reported ethnicity, biopsy of nail apparatus is highly unlikely to uncover invasive ALM in children younger than 12 years. Prospective studies are needed to understand how incidence of melanonychia, age, gender, and ethnicity/race influence parental and clinicians’ perceptions of melanoma risk.

## INTRODUCTION

There is limited epidemiological evidence in children regarding subungual melanoma (SUM), a form of cutaneous melanoma and type of acral lentiginous melanoma (ALM). Reported cases of pre-adolescent children diagnosed with SUM presenting as melanonychia remain controversial.^1-3^ Further, there are no widely accepted guidelines on who should undergo biopsy of the nail apparatus to screen for SUM. Notably, for children younger than 12 years living in the US, from 2000 to 2015 there are no reported cases of invasive ALM in the Surveillance, Epidemiology, and End Results (SEER) Program.^4^ The SEER Program collects cancer diagnoses, treatment, and survival for approximately 30% of the US population. However, ALM increases with age, from an estimated 0.1 per 1,000,000 person-years in adolescents (<20 years old) to 9.3 per 1,000,000 person-years in individuals 80 to 84 years old.^5^ These observed age differences are complicated by apparent dramatic ethnic differences in ALM prevalence and survival rates. For example, in the US, incidence of ALM is highest in Hispanic whites (2.5 per 1,000,000 person-years); however, Asian/Pacific Islanders reportedly have the lowest 10-year survival rates (54.1%).^5^ The aims of our study were to (1) Utilize an ethnically diverse pediatric cohort to determine how frequently children were diagnosed by a medical provider with melanonychia, (2) Characterize patients who underwent biopsy because of concern from SUM, and (3) Determine which, if any patients were diagnosed with melanoma.

## MATERIALS AND METHODS

We conducted a retrospective chart review of Rady Children’s Hospital San Diego’s (RCHSD) electronic health records (EHR) to identify patients diagnosed with melanonychia younger than 12 years, between January 1, 2010, and July 31, 2021. EPIC was used to search the over 2 million patients in our EHR using the SNOMED code 402633003 and the following SliceDicer™ search terms: “melanonychia”, “excision”, “melanonychia striata”, and “nail bed biopsy”. Additionally, EHR based searches were compared against records provided by the Department of Pathology at RCHSD. This study was approved by the University of California San Diego institutional review board.

## RESULTS

There were 623,805 patients younger than 12 years of age seen in the outpatient setting, of whom 204 were diagnosed with melanonychia. Average age was 7.2 years for melanonychia diagnosis, and 5.1 years (range 0.9-10.9) for those biopsied. Nail apparatus biopsies were performed in 22 different individuals and involved the hand 17/22 (77%) and the foot 5/22 (23%). A total of 4 individuals underwent a second biopsy of the same nail. For all individuals who underwent biopsy, none were diagnosed with melanoma or SUM. Pigment in an otherwise benign appearing biopsy was documented in 20/22 (91%) patients and histologic atypia in 2/22 (9%). Inadequate specimen sampling resulted in a second biopsy for 2 individuals. The presence of Hutchinson’s sign was documented in 9/22 (41%) patients. Whole nail involvement was documented in 3/22 (14%) patients. The average width for linear melanonychia was 2.3 mm for the 17 patients biopsied with a documented measurement. In patients who underwent biopsy, pigment recurrence was documented in 6/22 (27%), with this given as the reason for a second biopsy of the same nail in 2 patients. Males were diagnosed with melanonychia and underwent biopsy more often than females. Many of the biopsies were performed in Hispanic/Latino 11/22 (50%) and Asian/Pacific Islander 3/22 (14%) patients. For context, Hispanic/Latinos and Asian/Pacific Islanders represent respectively, 46% and 10% of San Diegans under age 19 years. All children who underwent nail biopsies are currently alive. Over the study period, six patients with melanoma received treatment at RCHSD, none involving the nail apparatus.

## DISCUSSION

Our cross-sectional study of over 2 million patient records from a regional pediatric healthcare system provides observational evidence that biopsy of the nail apparatus is unlikely to uncover invasive ALM in children younger than 12 years, regardless of reported ethnicity. Additionally, we found that regardless of a reassuring initial biopsy, pigment recurrence is common following biopsy, and its presence may prompt concern and result in re-biopsy. Use of EHR data is a limitation, we believe this underestimates prevalence of melanonychia in this population. However, a diagnosis of melanonychia in a patient’s record likely does reflect a perceived need for follow-up, and either provider or parental concern for malignancy. The absence of reliable estimates of melanonychia in populations made-up of children from diverse genetic ancestries limits use of inferential statistical analysis and the development of evidence-based recommendations for SUM screenings.

## CONCLUSION

It is not race but age that likely explains risk of SUM in children. Our findings are consistent with smaller prospective and retrospective observational studies, as well as the SEER Program cancer registry. Prospective studies are needed to understand how incidence of melanonychia, age, gender, and ethnicity/race influence parental and clinicians’ perceptions of melanoma risk and decision to biopsy.

## Data Availability

All data produced in the present study are available upon reasonable request to the authors.

## ACKNOWLEDGEMENTS

We would like to thank Dr. Denise Malicki MD and Dejon Welch (Division of Pathology at Rady Children’s Hospital San Diego) for completing SNOMED based searches of existing pathology reports and the Research Informatics team at Rady Children’s Hospital San Diego for performing EPIC SlicerDicer™ based data extractions.

---

**Table 1.** Characteristics of all patients diagnosed with melanonychia and biopsied patients.

| Patient Characteristic | Patients with Melanonychia (N=204) | Biopsied Patients (N=22) |
| --- | --- | --- |
| Mean age at diagnosis, years (Range) | 7.2 (0.67-11.96) | 4.2 (0.67-9.85) |
| Mean age at biopsy, years (Range) | N/A | 5.1 (0.90-10.90) |
| Sex, N (%) |  |  |
| Male | 117 (57) | 14 (64) |
| Female | 87 (43) | 8 (36) |
| Ethnicity/Race, N (%) |  |  |
| Hispanic or Latino | 100 (49) | 11 (50) |
| Asian and Pacific Islander | 34 (17) | 3 (14) |
| Black/African American | 17 (8) | 1 (4.5) |
| Non-Hispanic White | 27 (13) | 1 (4.5) |
| Other/Decline to Answer | 26 (13) | 6 (27) |

N/A, not applicable

